# Geographical disparities and differences in medical specialty prescribing of dronabinol in Medicare from 2014 to 2019

**DOI:** 10.1101/2022.07.20.22277818

**Authors:** Dayna S DeSalve, Kenneth L McCall, Brian J Piper

## Abstract

**Purpose:** The purpose of our study was to investigate dronabinol prescribing in Medicare from 2014 to 2019 by provider specialty and state.

**Methods:** Data was collected and analyzed from the Centers for Medicare & Medicaid Services databases from 2014 to 2019. The mean number of prescriptions for each area of practice, each individual year, and for 2014 to 2019 overall for the 50 United States and District of Columbia was determined. The prescriptions were separated by state and the state totals were determined. Individual states with dronabinol prescriptions ≥1.96 standard deviations (SD) from the mean were identified as significant.

**Results:** The total number of dronabinol prescriptions decreased 9.1% from 2014 to 2019. Dronabinol prescriptions were more concentrated in the eastern United States in 2019 than compared to 2014 [Tennessee (107.2), Kentucky (94.2), and West Virginia (87.6) (>1.96 SD)]. The largest portion of dronabinol prescriptions originated from primary care (1,736) compared to specialty areas of practice (1,233). Internal medicine (789.5), family medicine (608.8), hematology-oncology (343.3), nurse practitioners (337.3), and infectious disease (271.0) had the highest average number of dronabinol prescriptions per year (p<0.05). The areas of practice with the highest ratio of percent dronabinol prescriptions to percent Medicare utilization were infectious disease (15.8), hematology-oncology (12.2), and medical oncology (12.1).

**Conclusion:** Dronabinol usage declined among Medicare patients and became more concentrated in the eastern United States. Most prescriptions originated from primary care, although after accounting for Medicare patient utilization, the highest ratios originated from infectious disease, hematology-oncology, and medical oncology.

## Introduction

Dronabinol is a synthetic pharmaceutical-grade delta(Δ)9-tetrahydrocannabinol (THC) approved for treatment of chemotherapy associated nausea and vomiting unresponsive to conventional anti-emetics and acquired immunodeficiency syndrome (AIDS) associated anorexia.^1^ The first synthetic THC was approved by the United States (US) Food and Drug Association (FDA) in 1985 first as capsules, and recently as a solution in 2016.^2,3^ The newer form of dronabinol in oral solution is a schedule II drug, however, dronabinol in sesame oil in gelatin capsules is a schedule III drug.^3,4^ Dronabinol acts similar to *Cannabis sativa L*. (marijuana) and is a partial agonist at cannabinoid (CB) receptors in the brain, leading to appetite stimulation and the prevention of vomiting.^2,5^ THC/dronabinol binds CB_1_ and CB_2_ receptors, though CB_1_ are throughout the central nervous system and result in most of the psychoactive effects, while CB_2_ receptors are concentrated in the periphery.^6,7^ Along with suppressing nausea and vomiting, dronabinol can lead to additional effects (euphoria, drowsiness, sedation, somnolence) that might help provide relief for patients receiving chemotherapy.^7^

THC, and dronabinol, are mainly metabolized by cytochrome P450 (CYP) enzymes in the liver, including CYP2C9, CYP2C19, and CYP3A4.^8^ There are many common prescription drugs that are CYP inducers or inhibitors, such as ciprofloxacin, fluoxetine, bupropion, amiodarone, and verapamil, that can alter the metabolism of THC.^9^ Thus, it is important to take prescription drug interactions into consideration when prescribing dronabinol since 69.0% of US adults aged 40-79 used at least one prescription drug in the past 30 days, while 22.4% used at least five or more prescription drugs.^10^ Additionally, dronabinol has a large volume of distribution (approximately 10L/kg), which allows for 4-6 hours of psychoactive effects after the onset (0.5-1 hour).^7^ It is readily absorbed (90-95%), but its high lipid solubility and first-pass hepatic metabolism leads to only 10-20% reaching systemic circulation.^7^

The introduction of synthetic cannabinoids into the medical world has resulted in misuse and appearance on the black market.^11,12^ Synthetic cannabinoids, like dronabinol, have a higher association with morbidity and mortality when used as a drug of abuse compared to phytocannabinoids, the natural alternative.^5,12^ Synthetic cannabinoid overdose is more associated with agitation and cardiac dysrhythmia than marijuana.^11^ However, synthetic cannabinoids, including dronabinol and nabilone, are regulated by the FDA, whereas the only cannabis-derived drug product regulated is a prescription cannabidiol.^13^

Research into cannabinoids is still in its early phases. Current investigations focus on potentially expanding the use of dronabinol into obstructive sleep apnea, chronic pain, and substance abuse and withdrawal, however, there are no guideline supported indications for use in these areas, yet.^5^ Physicians, and in most states, physician assistants and nurse practitioners can all prescribe dronabinol but there is a lack of research into which areas of practice and which states are prescribing this cannabinoid the most.^14,15^ Oncology and palliative care may prescribe dronabinol to treat nausea and vomiting caused by chemotherapy, while infectious disease may prescribe dronabinol for anorexia associated with AIDS.^14^ Additionally, ongoing state legalization of medical and/or recreational marijuana may affect the number of dronabinol prescriptions, with accessibility to natural THC and less need for a synthetic version. Initiatives for the legalization of recreational marijuana passed in the fall of 2014 for Alaska, Oregon, and District of Columbia, in 2016 for California, Maine, Massachusetts, and Nevada, and in 2018 for Michigan and Vermont.^16,17^ Medical marijuana has become legalized in Maryland, Minnesota, and New York in 2014, Louisiana in 2015, Arkansas, Florida, North Dakota, Ohio, and Pennsylvania in 2016, West Virginia in 2017, and Missouri, Oklahoma, and Utah in 2018.^17^

Our objectives were to investigate which areas of practice were prescribing dronabinol and which states had the most dronabinol prescriptions and how these have changed from 2014 to 2019 among Medicare patients.

## Methods

### Procedures

Drug and provider data was collected for dronabinol from the Medicare Part D Prescribers by Provider and Drug database for 2014 to 2019.^18^ These years were chosen for analysis because they were the most recent years provided by the database. The mean number of prescriptions for each area of practice, each individual year, and for 2014 to 2019 overall for the 50 United States and District of Columbia was determined. The areas of practice were classified into primary care (internal medicine, family medicine, nurse practitioner, physician assistant, general practice, pediatric medicine) or specialty, and the totals were determined for each year. The prescriptions were separated by state and the state specific totals were determined for 2014 and 2019. Data on Medicare enrollment was collected from the Medicare Total Enrollment database.^19^ Information on Medicare utilization by area of practice was collected from the Medicare Physician, Non-Physician Practitioner and Supplier database for 2014 to 2019.^20^ Data on medical marijuana legalization by states was collected from ProCon.org and the National Conference of State Legislatures.^21,22^

### Data Analysis

The analyses were: (1) the total number of prescriptions and prescriptions per 1,000,000 Medicare enrollees for each year; (2) the total number of prescriptions from highest to lowest per state, corrected for the number of Medicare enrollees for 2014 and 2019 with state values outside of 1.96 (B) and 1.5 (A) standard deviations (SD) marked; (3) heat maps for total number of prescriptions per state, corrected for the number of Medicare enrollees for 2014 and 2019; (4) the percent change in number of dronabinol prescriptions per state from 2014 to 2019; (5) the number of dronabinol prescriptions in primary care versus specialty areas of practice for 2014 through 2019 and for each year alone; (6) the average total number of prescriptions for 2014 to 2019 for each area of practice and for each year alone (for visualization purposes, the areas of practice with an average above 30 prescriptions per year were included, consisting of 15 areas of practice) with a one-way ANOVA analysis for statistical significance (p<0.05) and variability expressed by the standard error of the mean (SEM); (7) the ratio of percentage of total number of dronabinol prescriptions to the percentage of total Medicare utilization in each area of practice; (8) the average cost per day of dronabinol for 2014 to 2019; (9) the percent of total dronabinol prescriptions originating from each area of practice in 2014 and 2019. The data analyses were completed using GraphPad Prism v 9.4.0. Figures were constructed using GraphPad Prism v 9.4.0 and Heatmapper.^23^

## Results

Overall, the number of prescriptions for dronabinol decreased 9.1% from 3,267 in 2014 to 2,969 in 2019 (Figure 1a). Dronabinol prescriptions per 1,000,000 Medicare enrollees decreased from 61.83% to 49.28% from 2014 to 2019 (Figure 1b).

**Figure 1.**
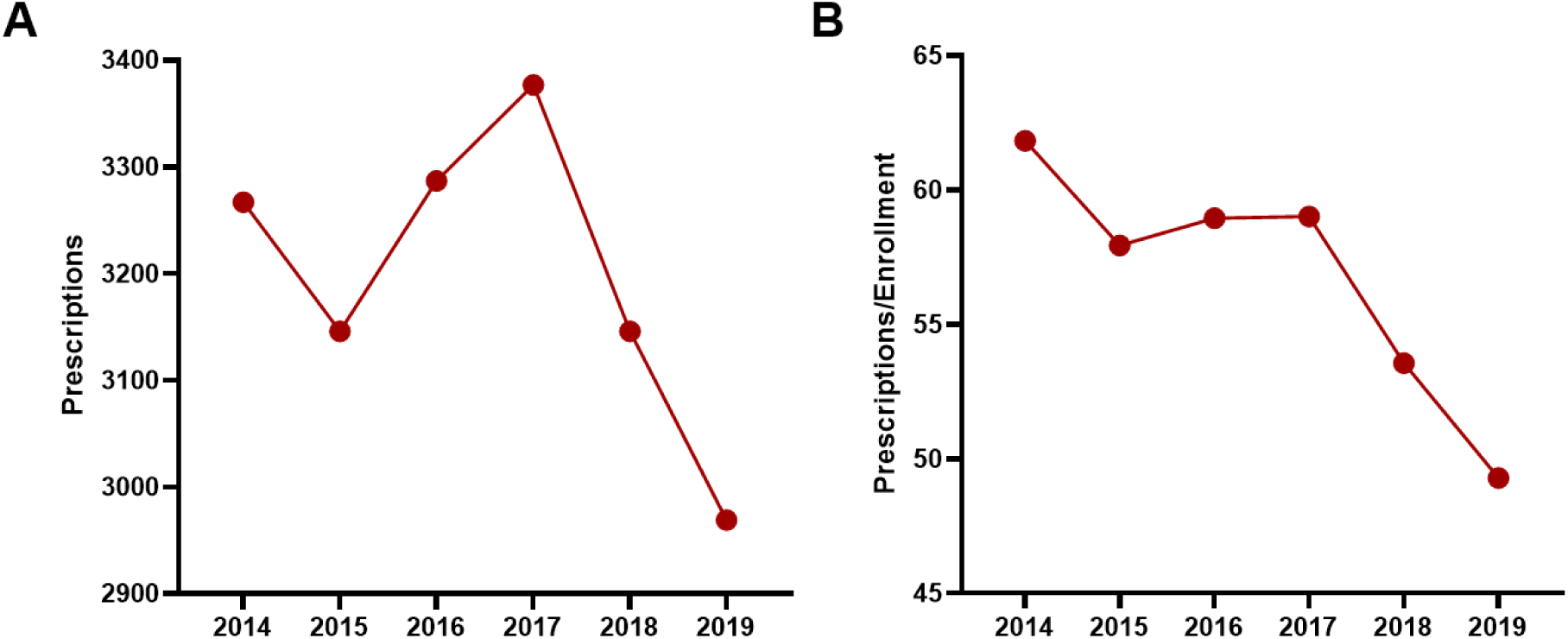
(A) Dronabinol prescriptions per year from 2014 to 2019. (B) Dronabinol prescriptions per 1,000,000 Medicare enrollees from 2014 to 2019.

Figure 2 shows the geographical distribution of dronabinol prescriptions in 2014 and 2019. Tennessee (107.2) had the highest number of dronabinol prescriptions in 2019, followed by Kentucky (94.2) and West Virginia (87.6) (>1.96 SD). Arizona (14.2) and Oregon (17.9) had the least number of dronabinol prescriptions in 2019 (<-1.5 SD). The highest number of dronabinol prescriptions in 2014 originated from Vermont (109.5), Tennessee (105.4), District of Columbia (104.6), and New Hampshire (101.6) (>1.96 SD). The lowest number of dronabinol prescriptions in 2014 originated from South Carolina (24.2) and Arizona (25.6) (<-1.5 SD). The ratio of highest to lowest number of dronabinol prescriptions per state for 2014 was 4.52 (Vermont:South Carolina), whereas in 2019, the ratio increased to 7.57 (Tennessee:Arizona).

**Figure 2.**
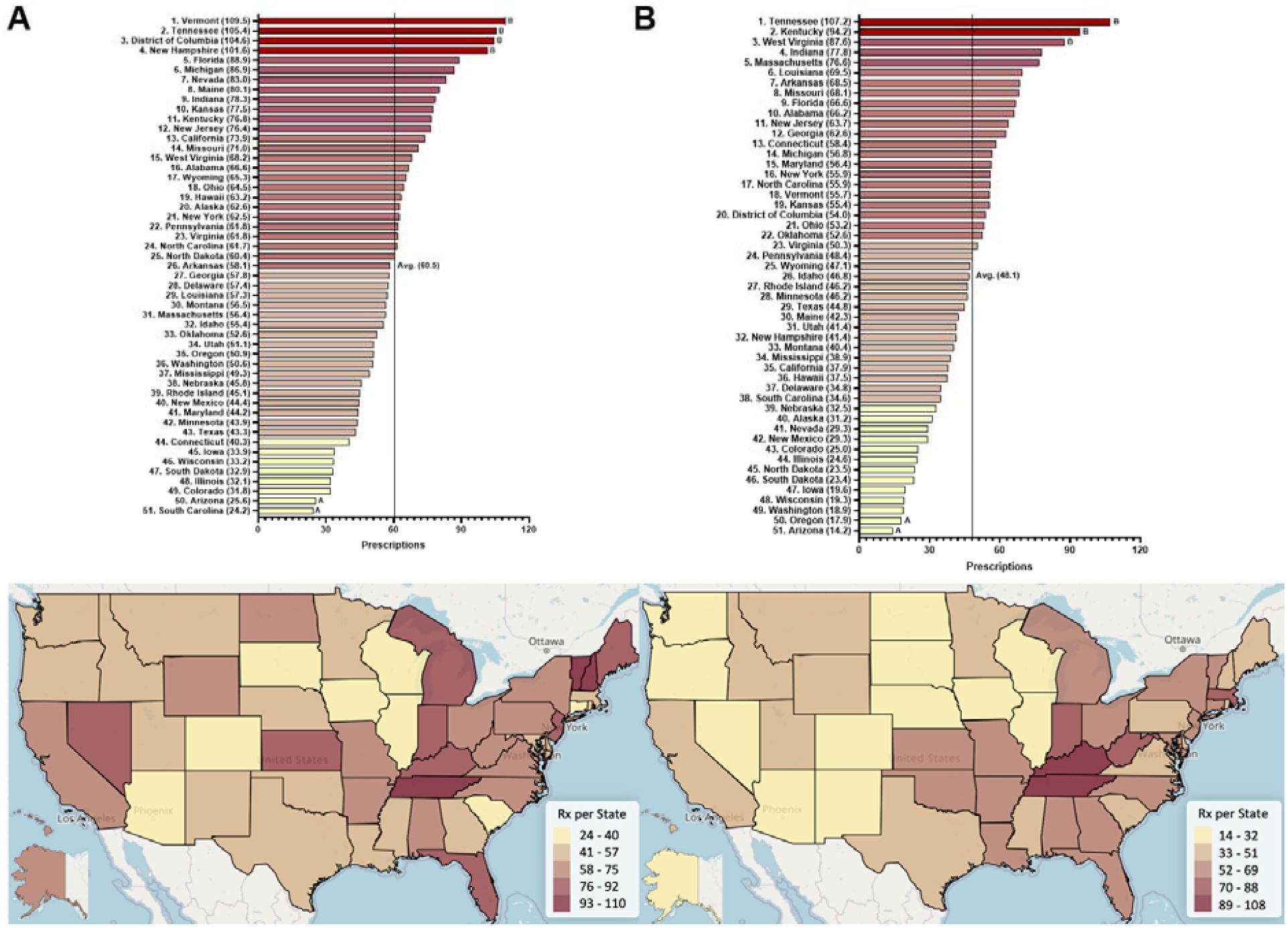
Dronabinol prescriptions per 1,000,000 Medicare enrollees. Ranked states (top) and heatmap (bottom) in 2014 (A) and 2019 (B). [States outside 1.96 SD (B) or 1.5 SD (A)]

Figure 3 compares the percent change in dronabinol prescriptions originating from each state from 2014 to 2019. Overall, over three-fifths (31 states and District of Columbia) decreased dronabinol prescribing from 2014 to 2019 while 18 states increased prescribing. Oregon (-146.7%), Nevada (-140.0%), and North Dakota (-133.3%) had the largest decrease in number of dronabinol prescriptions from 2014 to 2019. South Carolina (+38.9%), Connecticut (+35.9%), and Massachusetts (+33.0%) had the largest increase in number of dronabinol prescriptions from 2014 to 2019. Idaho was the only state that had the same number of dronabinol prescriptions for both 2014 and 2019.

**Figure 3.**
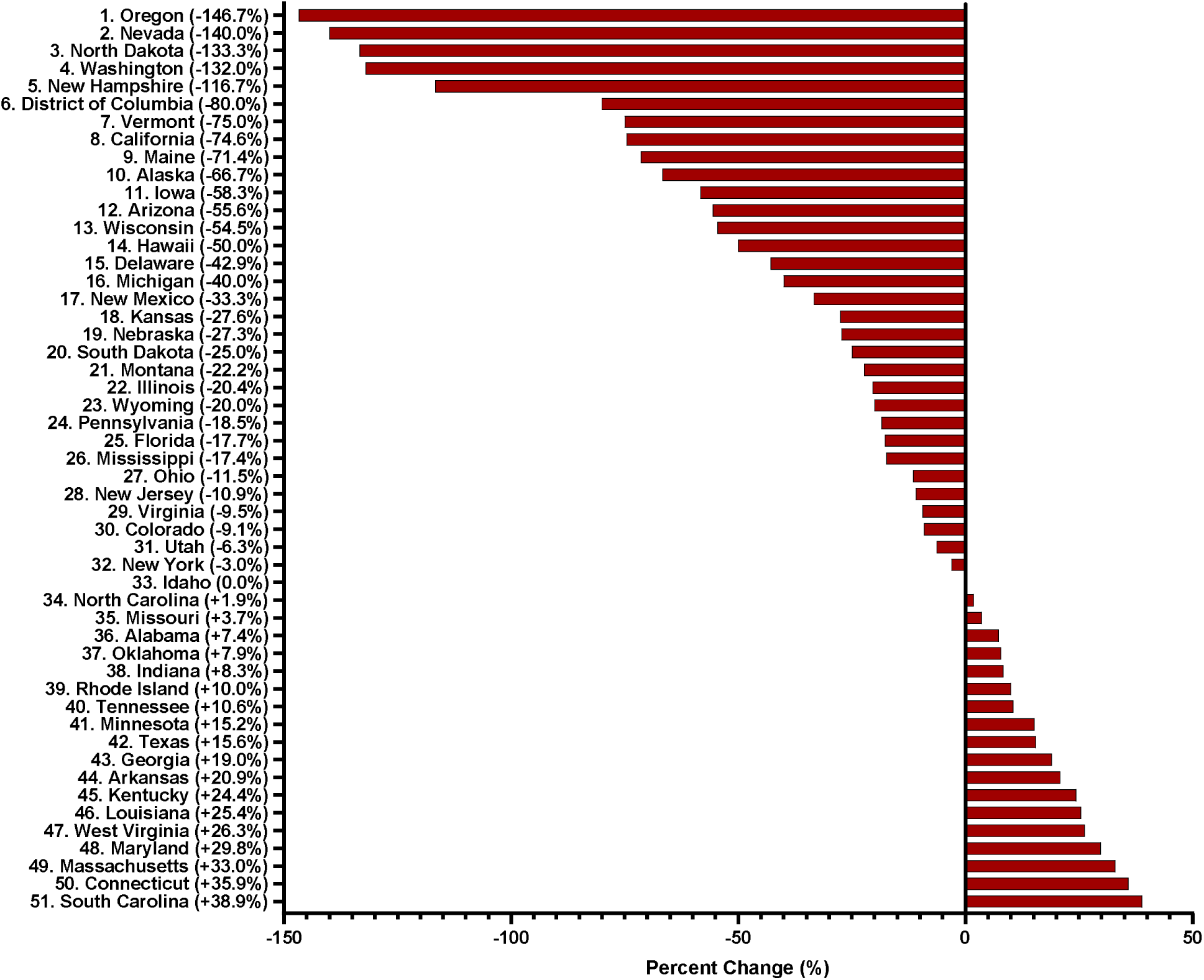
Percent change in number of dronabinol prescriptions originating from each state from 2014 to 2019.

Primary care constituted more than half of the yearly dronabinol prescriptions from 2014 to 2019, however, the percentage had decreased slightly from 61.4% to 58.5% (Figure 4). Figure 5 shows dronabinol prescriptions originating from each area of practice from 2014 to 2019. The areas of practice with the highest average number of dronabinol prescriptions from 2014 to 2019 were internal medicine (789.5) and family medicine (608.8), with hematology-oncology (343.3), nurse practitioners (337.3), and infectious disease (271.0) clustered next (p<0.05). Internal medicine (-29.0%), family medicine (-30.1%), and infectious disease (-26.9%) decreased in the amount of prescribing from 2014 to 2019. Hematology-oncology (+31.5%), nurse practitioner (+75.9%), and medical oncology (+62.0%) increased in the amount of prescribing from 2014 to 2019.

**Figure 4.**
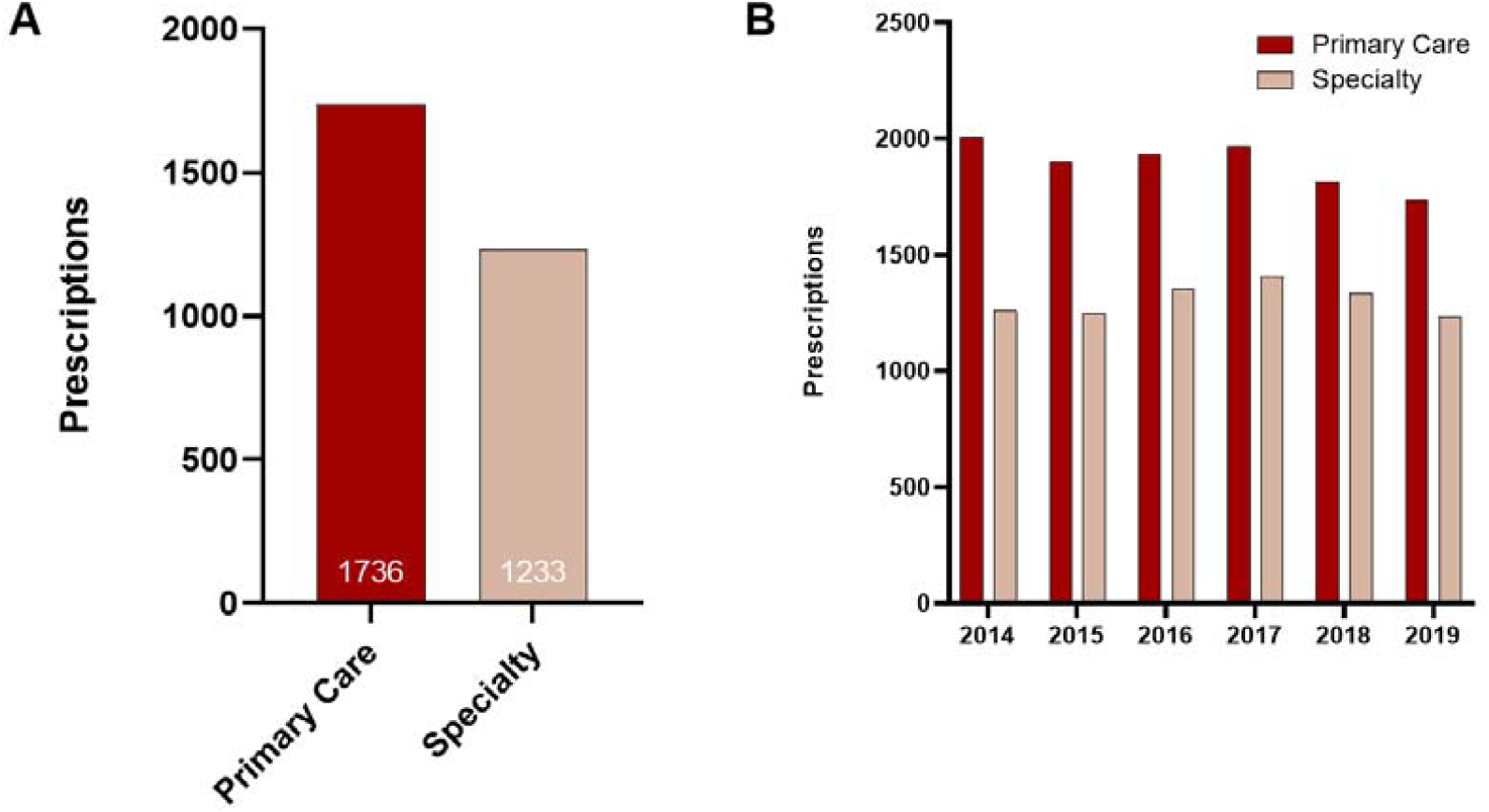
(A) Total number of dronabinol prescriptions in primary care and in specialty areas of practice from 2014 to 2019. (B) Total yearly number of dronabinol prescriptions in primary care and in specialty areas of practice each year from 2014 to 2019.

**Figure 5.**
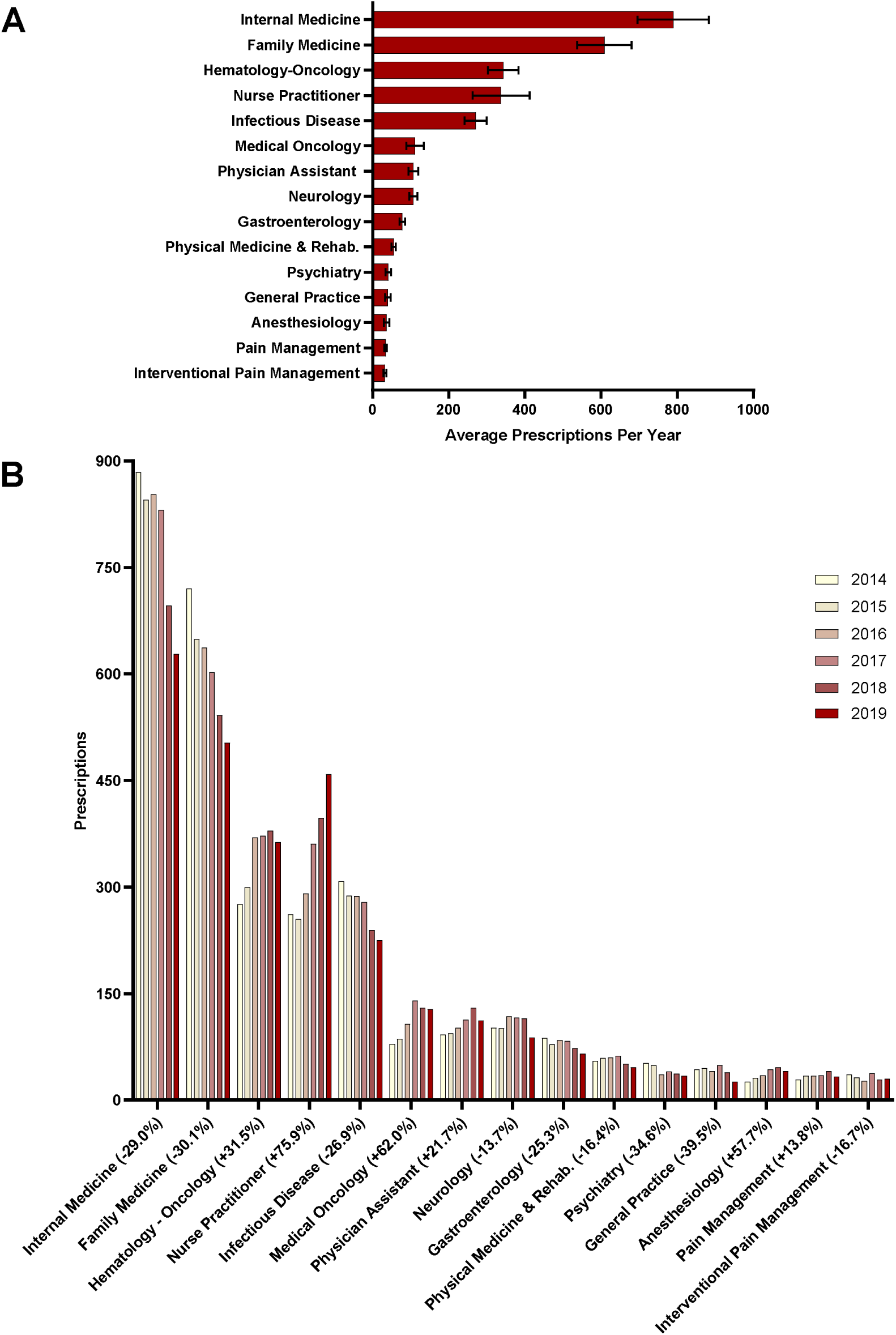
(A) Average number of dronabinol prescriptions in each area of practice from 2014 to 2019 with SEM bars. (B) Total dronabinol prescriptions in each area of practice per year from 2014 to 2019. Percent change from 2014 to 2019 noted in X axis labels.

Figure 6 shows the ratio of percent of total dronabinol prescriptions to percent of total Medicare utilization in 2019. Infectious disease (15.8), hematology-oncology (12.2), and medical oncology (12.1) had the highest ratios of percent of total dronabinol prescriptions to percent of total Medicare utilization in 2019. Gastroenterology (1.0) and physician assistant (1.0) had equal percentages. Anesthesiology (0.5) had the only ratio less than one.

**Figure 6.**
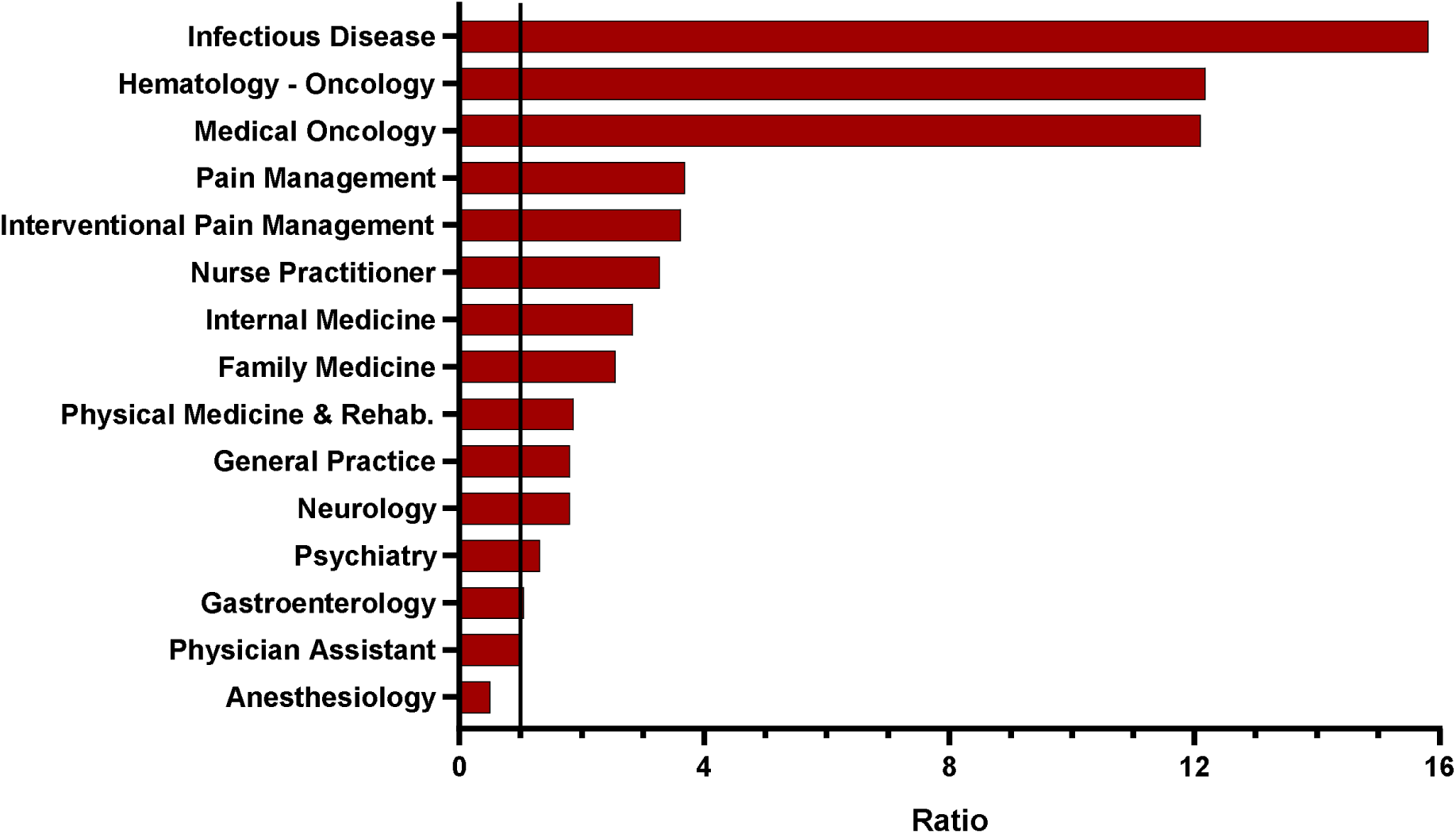
Ratio of percent of total dronabinol prescriptions to percent of total Medicare utilization in 2019. A ratio of one is marked with a vertical line.

## Discussion

Overall, the number of dronabinol prescriptions to Medicare patients decreased 9.1% from 2014 to 2019 while enrollment increased by 14.0%. There is inconsistency among states prescribing dronabinol, with a five-fold to eight-fold increase in the state level variation from 2014 to 2019. There is no good explanation for the geographical disparities, but we suspect it is due to state level differences in Medicare.

States with legalized medical marijuana, whether limited or full access, did not prove to have significant differences in the number of dronabinol prescriptions per 1,000,000 Medicare enrollees compared to states without any legalization of medical marijuana. It was hypothesized that the demand for dronabinol prescriptions was higher in states lacking medical and/or recreational marijuana legalization due to the absence of a similar alternative substance. In 2019, Tennessee (107.2) and Kentucky (94.2) had the largest number of prescriptions for dronabinol in the US. Both states only had limited access cannabis product laws in 2019: Tennessee only allowing low THC/high cannabidiol and Kentucky only allowing cannabidiol.^22^ Thus, legally obtaining medical and/or recreational marijuana in the state of Tennessee or Kentucky would not be an option for patients, leading to treatment with a synthetic form that requires a prescription, such as dronabinol.^22^ Certain states did have major changes in the number of dronabinol prescriptions from 2014 to 2019, which was hypothesized due to the passage of legislature that legalized medical and/or recreational marijuana. California and Nevada legalized the purchase of recreational marijuana in 2016, effective in 2016 and 2017 respectfully, which could explain why California dropped from number 13 to 35 (-74.6%) and Nevada from number 7 to 41 (-140.0%).^22,24-26^ North Dakota legalized medical marijuana in 2016 and dropped from number 25 to 45 (-133.3%).^22^ New Hampshire dropped from 4 to 32 (-116.7%) after legalizing the purchase of medical marijuana effective in 2016.^27^ Maine passed a ballot legalizing marijuana in 2016, effective in 2017, dropping rank from 8 to 30 (-71.4%).^26^ Oregon legalized marijuana late 2014, effective mid-2015, dropping from 35 to 50 (-146.7%).^26^ The states that have legalized medical and/or recreational marijuana may see a decrease in dronabinol or cannabinoid prescriptions in general due to increased accessibility to non-synthetic forms.

Compared to specialty areas of practice, primary care accounted for more than half of the prescriptions of dronabinol each year, with an average of 59.2% from 2014 to 2019. Internal medicine (789.5) and family medicine (608.8) had the highest average number of dronabinol prescriptions from 2014 to 2019, followed by hematology-oncology (343.3), nurse practitioner (337.3), and infectious disease (271). All of these values were proved to be statistically significant using a t-test (two sample assuming unequal variances). However, in 2019, primary care accounted for only 32.8% of Medicare utilization. The areas of practice with the highest percentage of total Medicare utilization in 2019 were internal medicine (7.98%), family medicine (7.11%), and nurse practitioner (5.04%). The same areas of practice had the highest percentage of total dronabinol prescriptions [internal medicine (22.58%), family medicine (18.09%), nurse practitioner (16.50%)]. However, infectious disease (15.8), hematology-oncology (12.2), and medical oncology (12.1) had the three highest ratios of percent total dronabinol prescriptions to percent total Medicare utilization in 2019. Thus, for the number of patients who utilized these specialty areas (infectious disease, hematology-oncology, medical oncology) the largest numbers of dronabinol prescriptions originated. This is supported by the fact that the two approved uses of dronabinol are AIDS associated anorexia (infectious disease) and chemotherapy associated nausea and vomiting (oncology).^1^ Internal medicine and family medicine experienced the largest decrease in relative prescribing, which is likely due to many patients no longer seeking a prescription cannabinoid from a general practitioner when they can now acquire herbal cannabis. Whereas the specialties of hematology-oncology and medical oncology experienced an increase in relative prescribing. The increase in relative prescribing of dronabinol by nurse practitioners is likely due to more states allowing nurse practitioners and/or more nurse practitioners obtaining a Drug Enforcement Administration (DEA) number to prescribe controlled substances as their scope of practice increases.

## Limitations

A strength of this study is the sample included Medicare patients who are over 65 years of age or disabled and who are at greatest likelihood of polypharmacy and experiencing drug interactions. Nurse practitioner and physician assistant were provider types listed in the database Medicare Part D Prescribers – by Provider and Drug. Follow up with the Centers for Medicare and Medicaid Services provided no additional insight or detail on these types and which departments they originated from.

## Conclusion

Dronabinol usage declined among Medicare patients, which may be related to legalization of medical and/or recreational marijuana, as it offers an alternative and natural source of THC. There is inconsistency among states prescribing dronabinol, with a five-fold to eight-fold increase in the state level variation. Most prescriptions originate from primary care areas of practice, although when accounting for Medicare patient utilization, the highest ratios of prescriptions to utilization originate from infectious disease, hematology-oncology, and medical oncology.

## Data Availability

All data produced in the present study are available upon reasonable request to the authors and are available online at Centers for Medicare and Medicaid Services.

https://data.cms.gov/provider-summary-by-type-of-service/medicare-part-d-prescribers/medicare-part-d-prescribers-by-provider-and-drug

https://data.cms.gov/summary-statistics-on-beneficiary-enrollment/medicare-and-medicaid-reports/medicare-total-enrollment

https://data.cms.gov/summary-statistics-on-use-and-payments/medicare-service-type-reports/medicare-physician-non-physician-practitioner-supplier

## Ethics Statement

Procedures were approved by the Institutional Review Board of Geisinger (IRB: 2022-0533). Brian J Piper was supported by the Health Resources Services Administration (D34HP31025) and the Geisinger Academic Clinical Research Center.

## Acknowledgements

Iris Johnston for inter-library loans.

