## Supplemental Table 1 for "Geographical disparities and differences in medical specialty prescribing of dronabinol in Medicare from 2014 to 2019"

**Supplemental Table 1.** Percentage of total Medicare utilization and percentage of total dronabinol prescriptions in 2019 with expression as a ratio of percent of total dronabinol prescriptions to percent of total Medicare utilization for each area of practice.

|  | Medicare Utilization (%) | Dronabinol Prescriptions (%) | Ratio |
| --- | --- | --- | --- |
| Infectious Disease | 0.51 | 8.09 | 15.82 |
| Hematology-Oncology | 1.07 | 13.05 | 12.18 |
| Medical Oncology | 0.38 | 4.60 | 12.10 |
| Pain Management | 0.32 | 1.19 | 3.69 |
| Interventional Pain Management | 0.30 | 1.08 | 3.61 |
| Nurse Practitioner | 5.04 | 16.50 | 3.27 |
| Internal Medicine | 7.98 | 22.58 | 2.83 |
| Family Medicine | 7.11 | 18.09 | 2.54 |
| Physical Medicine and Rehab. | 0.89 | 1.65 | 1.86 |
| General Practice | 0.52 | 0.93 | 1.81 |
| Neurology | 1.75 | 3.16 | 1.81 |
| Psychiatry | 0.93 | 1.22 | 1.31 |
| Gastroenterology | 2.23 | 2.34 | 1.05 |
| Physician Assistant | 3.95 | 4.03 | 1.02 |
| Anesthesiology | 2.93 | 1.47 | 0.50 |
