## Supplemental Figure 2 for "Geographical disparities and differences in medical specialty prescribing of dronabinol in Medicare from 2014 to 2019"

**Supplemental Figure 2**. Percent of total dronabinol prescriptions in each area of practice in 2014 and 2019. Percent change from 2014 to 2019 noted in X axis labels.

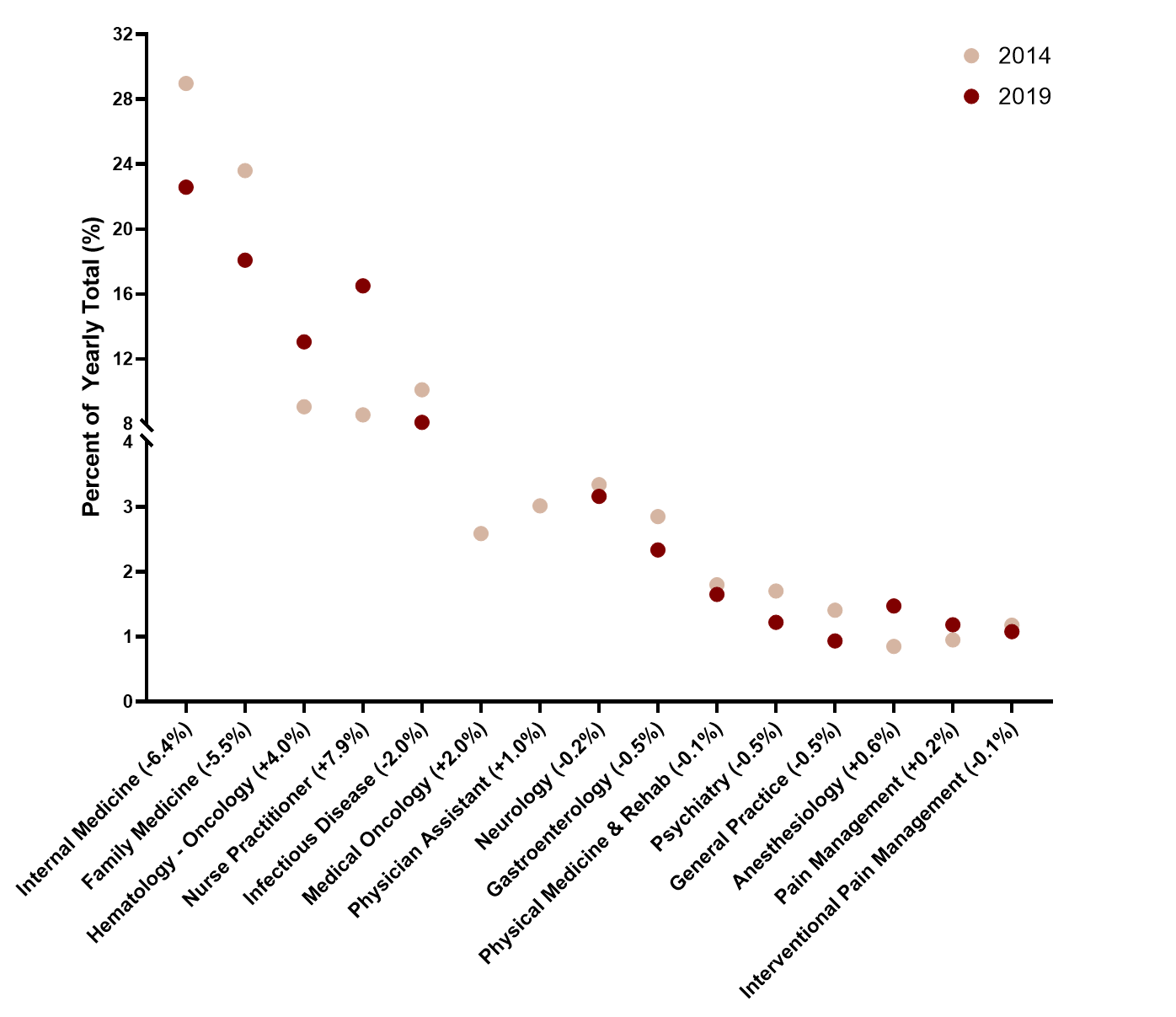
