## Supplemental Figure 1 for "Geographical disparities and differences in medical specialty prescribing of dronabinol in Medicare from 2014 to 2019"

**Supplemental Figure 1.** Average cost per day (±SEM) of prescribed dronabinol to Medicare patients.

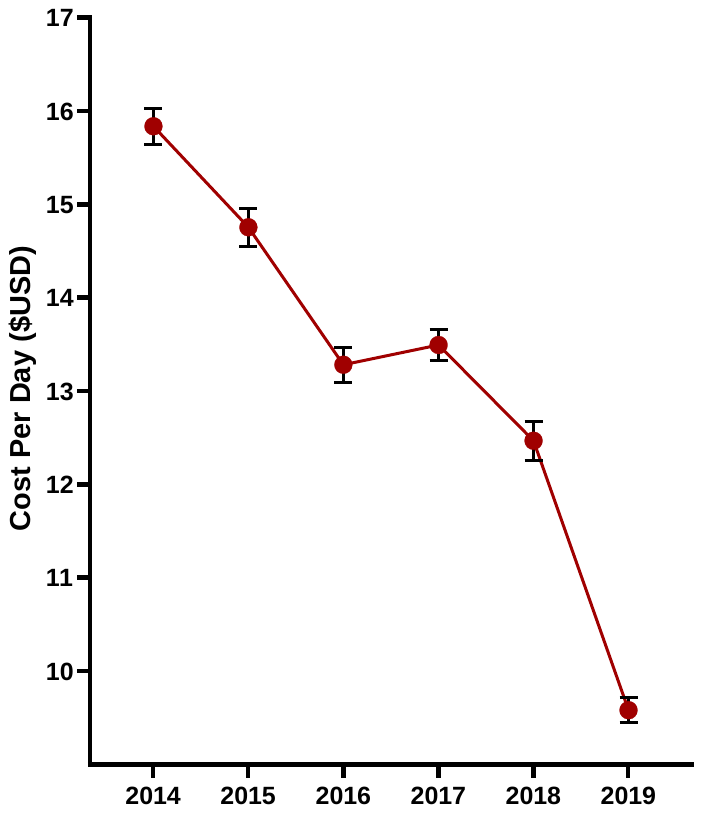

Average daily cost of prescribed dronabinol decreased 39.5% from $15.84/day in 2014 to $9.58/day in 2019.
